# Evaluating the serological status of COVID-19 patients using an indirect immunofluorescent assay, France

**DOI:** 10.1101/2020.05.05.20092064

**Authors:** S. Edouard, P. Colson, C. Melenotte, F. De Pinto, L. Thomas, B. La Scola, M. Million, H. Tissot-Dupont, P. Gautret, A. Stein, P. Brouqui, P. Parola, J.-C. Lagier, D. Raoult, M. Drancourt

**Affiliations:** IHU; Aix Marseille Universite-IHU Mediterranee Infection

## Abstract

An indirect immunofluorescent assay was developed in order to assess the serological status of 888 RT-PCR-confirmed COVID-19 patients (1,302 serum samples) and controls in Marseille, France. Incorporating an inactivated clinical SARS CoV-2 isolate as the antigen, the specificity of the assay was measured as 100% for IgA titre ≥ 1:200; 98.6% for IgM titre ≥ 1:200; and 96.3% for IgG titre ≥ 1:100 after testing a series of negative controls as well as 150 serums collected from patients with non-SARS-CoV-2 Coronavirus infection, non-Coronavirus pneumonia and infections known to elicit false-positive serology. Seroprevalence was then measured at 3% before a five-day evolution up to 47% after more than 15 days of evolution. We observed that the seroprevalence as well as the titre of specific antibodies were both significantly higher in patients with a poor clinical outcome than in patients with a favourable evolution. These data, which have to be integrated into the ongoing understanding of the immunological phase of the infection, suggest that serotherapy may not be a therapeutic option in patients with severe COVID-19 infection. The IFA assay reported here is useful for monitoring SARS-CoV-2 exposure at the individual and population levels.

## INTRODUCTION

The SARS-CoV-2 is a coronavirus belonging to the genus *Betacoronavirus* that emerged in humans in December 2019 (1). It was first described in China before spreading and being classified as a pandemic (2). It causes a respiratory disease known as Covid-19 that is usually mild but can result in a severe and even life-threatening pneumonia, particularly in elderly people (2, 3). On 24 April 2020, 2,699,338 SARS-CoV-2 infections and 188,437 associated deaths had been reported worldwide [https://coronavirus.jhu.edu/map.html].

To date, the virological diagnosis of infections by SARS-CoV-2 has been essentially based on real-time reverse transcription PCR (4). This virus has been shown to elicit specific antibodies during the course of infection (1, 5). This serological response has mainly been analysed using enzyme-linked or chemiluminescence immunoassays among exposed populations in China and neighbouring countries. Previous studies showed that specific IgG, IgM and IgA were produced in response to the infection (6). The kinetics of these three classes of antibodies have been described, yet correlations with the clinical outcome of the patients has been poorly reported (6).

In this study, we implemented an indirect immunofluorescent assay for the detection of anti-SARS-CoV-2 antibodies, and observed significant differences in the seroprevalence and antibody titres between groups of patients depending on their clinical outcome.

## PATIENTS AND METHODS

### Study design

A cohort of patients with confirmed SARS-CoV-2 infection was studied at the Institut Hospitalo-Universitaire (IHU) Méditerranée Infection in Marseille, France, as previously described (7). All patients presenting symptoms compatible with COVID-19 and contacts of suspected and confirmed COVID-19 cases were tested using a SARS-CoV-2 specific qRT-PCR assay (7, 8). Treatment with hydroxychloroquine (HCQ) associated with azithromycin (AZ) was proposed to all qPCR-positive patients who enrolled on a voluntary basis if they did not present contraindications (7). Patients were followed-up on an out-patient basis at our day care hospital or were hospitalised in the infectious disease units of the IHU, in intensive care units or in other medical departments of the Assistance Publique-Hôpitaux de Marseille, depending on the severity of the disease. We included in the present study all patients from the previous study by Million *et al*. for whom ≥1 serum sample was available for serological testing as part of the routine care of these patients. The serum samples were tested retrospectively using an indirect immunofluorescence assay (IFA). The time of serum collection was determined relative to the date of the onset of symptoms. The non-interventional nature of this study was approved by the Ethical Committee of the IHU Méditerranée Infection under no. 2020-13.

### Case definition

SARS-CoV-2 infection was defined by clinical, radiological, and microbiological criteria as previously reported (3, 7). Briefly, the national early warning score (NEWS) for COVID-19 was used for the classification of clinical presentation of patients. Virological evidence of the infection was based on a positive qRT-PCR on a nasopharyngeal sample or another respiratory sample. Pulmonary involvement was evaluated by chest low-dose computed tomography for all patients. Five groups of patients were constituted according to the following criteria (7): (1) Patients with mild disease and good clinical and virological outcome (GO; n= 681); (2) Patients with poor virological outcome defined by persistence at day 10 or more of viral detection in respiratory samples (PVirO; n= 100); (3) Patients who received HCQ + AZ treatment for more than three days, with poor clinical outcome requiring prolonged hospitalisation for 10 days or more despite three days or more of HCQ + AZ treatment (PClinO1; n= 53); (4) Patients who received HCQ + AZ treatment for fewer than three days, with poor clinical outcome requiring prolonged hospitalisation for 10 days or more (PClinO2; n = 25); (5) Patients with poor clinical outcome requiring prolonged hospitalisation for 10 days or more leading to death (PClinO3; n= 29). Main characteristics of the patients in each group are summarised in Table 1.

### Indirect immunofluorescence assay

Anti-SARS-Cov 2 antibodies were detected using an in house indirect immunofluorescence assay (IFA), as previously described (9). Vero E6 cells (ATCC CRL-1586, Rockville, MD, USA) infected with the SARS-CoV2 strain IHU-MI2 (full genome sequence of this strain was deposited under the European Molecular Biology Laboratory EMBL project accession no. PRJEB38023) (10) were harvested between 24 hours and 48 hours post-inoculation when cytopathic effect begins to be observed before massive cell lyses begin, washed with sterile phosphate buffered saline (PBS) (Oxoid, Dardilly, France) and inactivated using 5% paraformaldehyde. This preparation was used as the antigen and 50 nL of antigen were spotted on each well of 18-well microscope glass slides using Echo 525 Liquid Handler instruments (Labcytes, Cannock, United Kingdom) that uses acoustic energy to transfer liquid from a 96-well plate containing the antigen to slides. Fifty nanolitres of uninfected Vero cells were also spotted on each well as a negative control and a clinical isolate of *Staphylococcus aureus* (identified by matrix-assisted laser desorption ionization-time of flight mass spectrometry) (11) was spotted on each well in order to ensure further serum deposition, as previously described (12). Each slide was air dried, fixed in acetone for 10 minutes and conserved at 4°C in the dark.

In a first step, each serum sample was screened for the presence of anti-SARS CoV-2 antibodies using the IFA, as previously described (9). Serum samples were heat-decomplemented for 30 minutes at 56°C, diluted in 3% PBS-milk and 25 μL of a 1:50 dilution and a 1:100 dilution were pipetted onto a 18-spot slide then incubated for 30 minutes at 37°C in the dark to be screened for the detection of total immunoglobulin (IgT). After washing thrice, the slides with sterile PBS for 10 minutes, 25 μL of total FITC-conjugated IgT anti-human immunoglobulin (Bio-Rad, Marnes-la-Coquette, France) with 0.5% Evans blue (Bio-Rad) were incubated for 30 minutes at 37°C. After washing, slides were observed under a fluorescence microscope (AxioSkop 40, Zeiss, Marly le Roi, France). In a second step, all the serum samples screened positive at a 1:100 dilution were quantified for IgG, IgM and IgA as reported above, except that serum samples were diluted up to 1:1,600 for IgA and IgM and 1:3,200 for IgG; and anti-IgG, anti-IgM and anti-IgA conjugates were used (bioRad). Serum samples exhibiting positivity at 1:3,200 were further tested up to 1:6,400. A serum sample exhibiting a 1:400 titre collected from one patient who was positive by SARS COV-2 RT-PCR, was anonymised and used as a positive control on each slide for screening and on each run for antibody quantification. A negative serum collected in December 2019 from a patient and PBS-milk 3% were used as negative controls on each slide screened. In order to interpret the IFA, any serum sample exhibiting IgG 1:100 was considered as positive; as well as any serum sample exhibiting isolated IgM or IgA 1:200.

### Serum samples

The specificity of the IFA was evaluated by testing four series of serum samples. Negative control samples (n = 200) had been collected from patients between November and December 2018 (before the COVID-19 epidemics in France). Further, serum samples known to be associated with nonspecific serological interference were collected from 14 patients diagnosed with Epstein-Barr virus infection; eight patients diagnosed with Cytomegalovirus infection; seven patients diagnosed with A hepatitis virus infection; 10 patients diagnosed with toxoplasmosis and 25 patients diagnosed with E hepatitis virus infection. Serum samples were also collected from 50 patients diagnosed with Coronavirus NL63, OC43, 229E or HKU1; as well as 36 sera collected from patients diagnosed with non-coronavirus pneumonia, including 14 *Mycoplasma pneumoniae* infections, 10 *Legionella pneumophila* infections, and 12 *Chlamydia pneumoniae* infections, in order to assess for potential cross-reactivity.

### Statistical analysis

To avoid bias in data analysis, we studied the serological response according to the time of sampling of the sera related to the date of the onset of symptoms. The analysis of sera was divided into different times (D0-D5, D6-D10, D11-D15 and D16-D38). For the studied of seroprevalence and for the comparison of IgG titre, we considered only the sera with the higher IgG titre or with the higher IgM or IgA titre when several sera were available for a same patient. For the data comparisons and statistical analyses, Fisher’s exact test or the Chi-squared test and standard statistical software (GraphPad Prism 5) were used. A p-value < 0.05 was considered statistically significant. ROC curves were calculated using GraphPad Prism 5.

## RESULTS

### IFI assay

In the negative control group of 200 serum samples collected from patients in November and December 2018 before the emergence of COVID-19 in France, no IgG and no IgA were detected and three samples exhibited a IgM titre of 1:25 for two samples and 1:100 for one sample (Figure 1). In the group of serum samples known to yield cross-reactivities, two samples collected from patients diagnosed with Epstein-Barr virus infection (n = 14) exhibited IgG titre 1:200, one to 1:100 and two IgM titre ≥ 1:100; two samples collected from patients diagnosed with Cytomegalovirus infection (n = 08) exhibited IgM titre 1:100; one sample collected from patients diagnosed with hepatitis A (n = 07) exhibited an IgM titre at 1:200 and two at 1:100; and one sample collected from patients diagnosed with hepatitis E (n = 25) exhibited IgG titre at 1:400; and one serum exhibited IgM titre at 1:100. Of the 50 serum samples collected from patients diagnosed with another Coronavirus other than COVID-19, none reacted in IgG, none reacted in IgA and six reacted at 1:100 in IgM, two reacted at 1:200 and one reacted at 1:800 (Table 2). No positivity was observed from 10 serum samples drawn from toxoplasmosis patients. Also, 36 sera collected from patients diagnosed with non-Coronavirus pneumonia yielded an IgG titre at 1:400 (n = 3) and an IgG titre at 1:100 (n = 6). Overall, 13/350 serum samples yielded a false positivity of IgG ≥ 1:100, yielding a 96.3% specificity for IgG; and 05/350 serum samples yielded a false positivity of IgM ≥ 1:200, yielding a specificity of 98.6% for IgM. Specificity of IgA titre of 1:200 was 100%.

**Figure 1.**
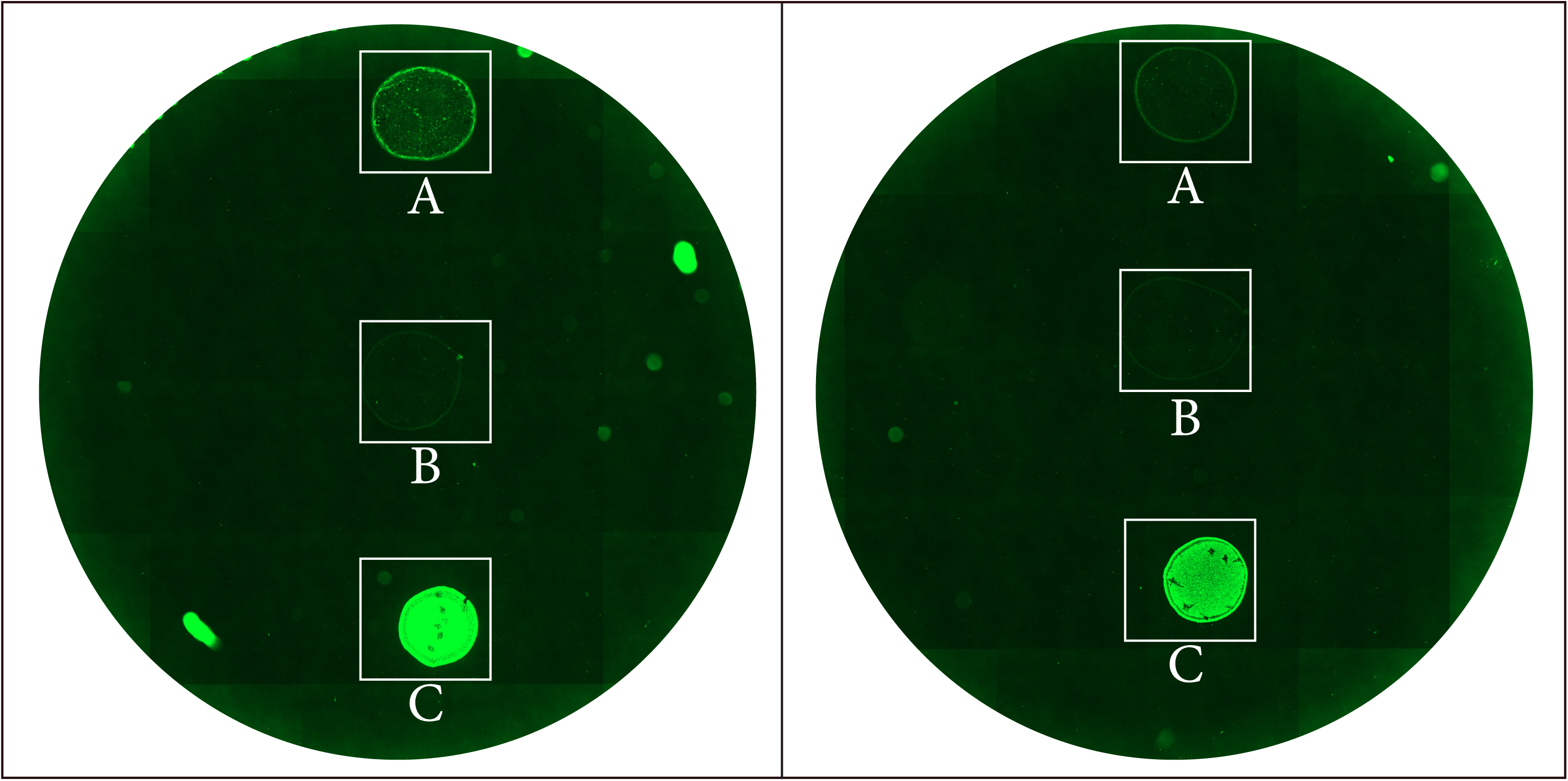
Picture of immunofluorescence assay of serum sample from a COVID-19 Infected patient. Each well of glass slides was spotted with SARS-Cov-2 antigen (A), non-infected VERO cells (B) and S. *aureus* antigen (C). Left panel, patient’s serum with anti-SARS-CoV-2 total immunoglobulins detectable at dilution 1:100. Patient presented IgG titer at 1:400, IgM titer at 1:50 and IgA titer at 1:100. Right panel, negative control serum. Slides were observed using Zeiss microscope, objective x40.

We then evaluated the serological response in a collection of 1,302 serum samples from 888 patients infected with SARS-CoV-2 between 12 March and 17 April 2020 (7). This cohort, which included 408 men (46%), had a median age of 45 years (range, 14–97 years). Median age of patients from PClinO1, PClinO2, PClinO3 group were significantly higher than the median age of patients from PVirO and GO group (p<0.0001). Serum samples had been collected at a median time of 15 days (range, 0–38 days) after onset of symptoms. Seventy (5.4%) sera were collected between D0-D5, 238 (18.3%) between D6-D10, 395 (30.3%) between D11-D15 and 599 (46%) between D15-D38. Multiple sera were available for 299 patients. At least one positive serology was found in 330 patients, leading to a global seroprevalence of 37.2%. The time distribution of positive serum samples was as follows: 3% between D0-D5, 13% between D6–D10, 27% between day D11–D15 and 47% after D16. We observed 88 (29%) seroconversions that occurred between D6–D10 in 6 (7%) cases, between D11–D15 in 25 (28%) cases and after D16 in 57 (65%) cases. Only two patients were observed to be positive within five days after onset of the illness, one patient exhibited IgG titre 1:100 and another patient with IgG titre at 1:1,600 and IgA at 1:100.

Detailing the results for each group of patients, the median time of serum sampling was 8, 11, 11, 16 and 16 days after the onset of symptoms for PClinO3, PClinO2, PClinO1, PVirO and GO, respectively. Global seroprevalence by group was 29% in PClinO3, 56% in PClinO2, 49% in PClinO1, 44% in PVirO and 29% in GO patients. Higher seroprevalence was observed in group of PClinO3, PClinO2, PClinO1 compared to GO group between D6-D10 but this was not significant. However, significant higher seroprevalence between patients with poor clinical outcome compared to patients with good clinical outcome was observed after D10 (Figure 2). Higher seroprevalence was found in PClinO3 (70%), PClinO2 (71%), PClinO1 (57%) compared to patients with good clinical outcome (GO) (37%), p=0.046, p=0.01 and p= 0.015, respectively. In particular, the five deda patients had exhibited positive serology after day 16. No significant difference was observed between PVirO and GO group.

**Figure 2.**
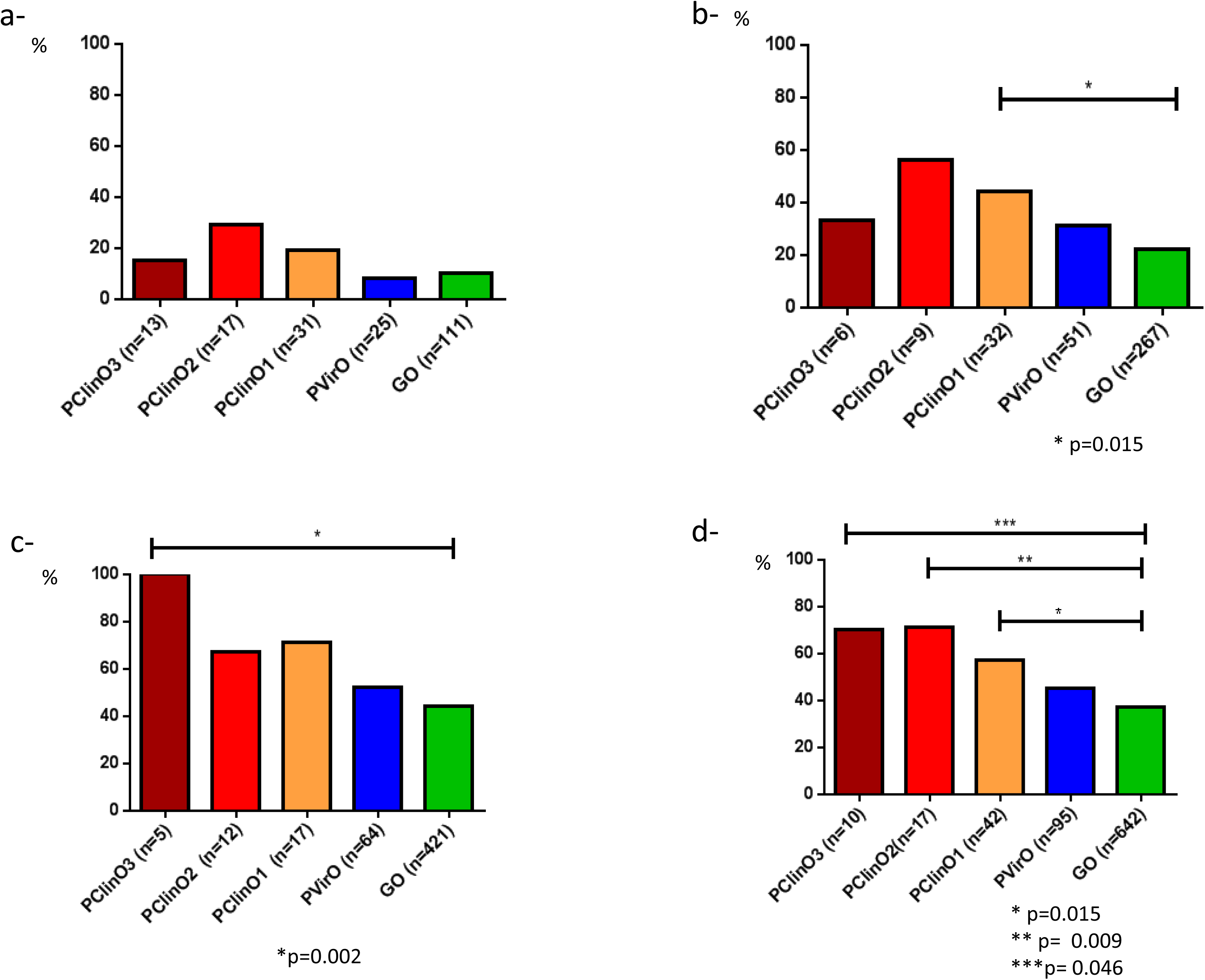
Comparison of seroprevalence among the five groups of patients (a) Between days 6 and 10 (b) Between days 11 and 15 (c) between days 16 and 38 (d) After day 38.

We also compared IgG titre between the five groups of patients on sera collected at least 10 days after the onset of symptoms. We found significant higher IgG titre in patients with a poor clinical outcome (died PClinO3, PClinO2, PClinO1) compared to patients with good outcome (GO) (p=0.0007) (Figure 3).

**Figure 3.**
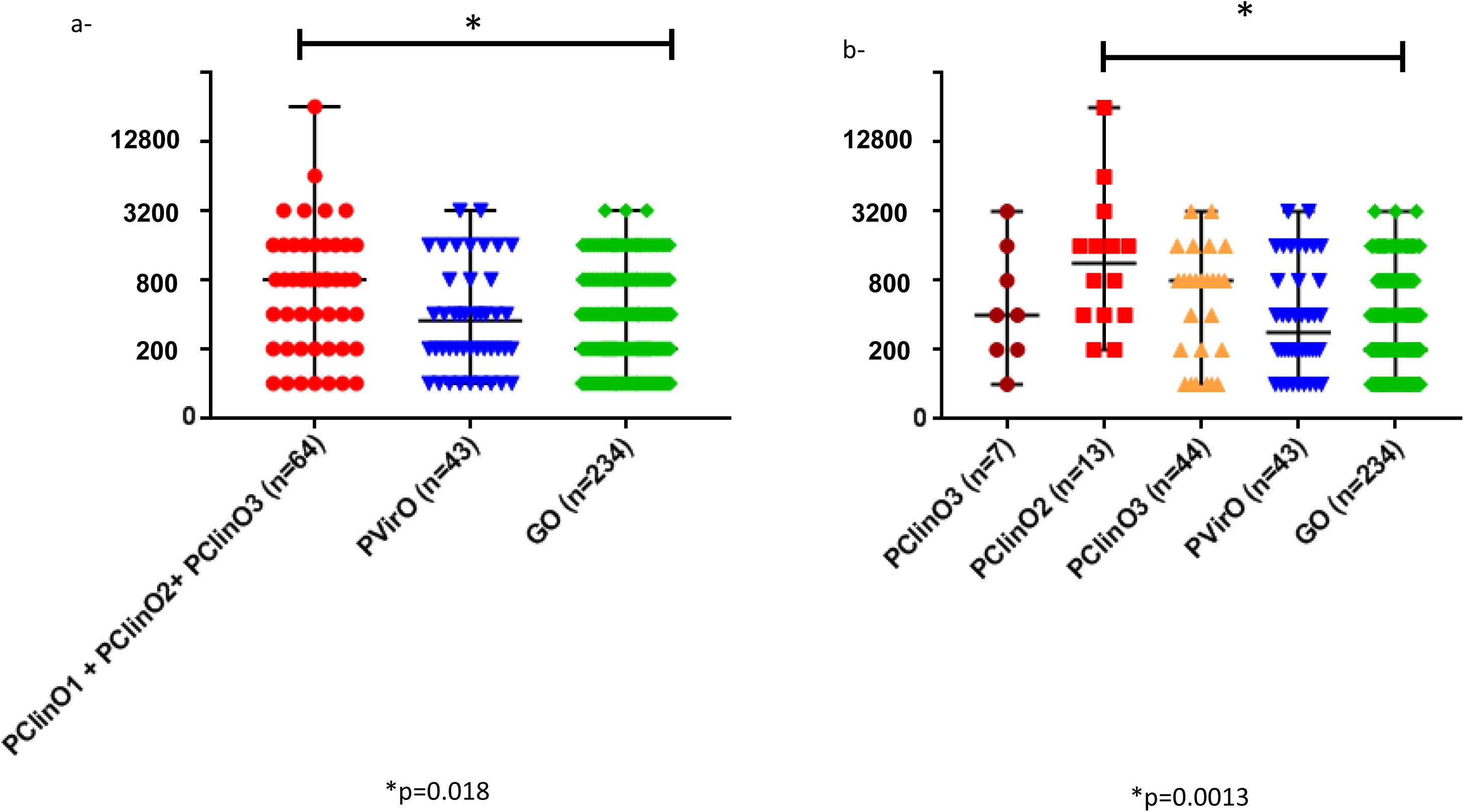
Comparison of IgG titre detected at least 10 days after the onset of symptoms between the different group of patients infected with SARS-CoV-2. When multiple sera were available for a same patient, only the sera with higher IgG titre were considered for this analysis.

## DISCUSSION

We developed an indirect immunofluorescence assay for the detection of IgG, IgM and IgA anti-SARS CoV-2 antibodies and we used it to assess the serological status of hundreds of COVID-19 patients and controls, as such an assay has been only reported on a very small group of patients (13). In order to avoid false negative results, the assay incorporated S. *aureus* as a control of deposition of tested sera, as *S*. *aureus* protein A and protein M bind non-specifically to any serum antibody (12). The assay also incorporated non-infected Vero cells on which the viral antigen has been produced, in order to identify false positive reactivities. Reading of both controls was incorporated into the interpretation algorithm. Accordingly, the specificity of the assay was measured at 100% for IgA, 98.5% for IgM and 95.9% for IgG.

Using this assay, we observed low values of seroprevalence, at 37% in RT-PCR confirmed COVID-19 patients, ranging precisely from 3% before five days’ evolution to 47% after 15 days’ evolution. However, seroconversions of specific IgM and IgG antibodies were observed as early as day four after the onset of symptoms, as previously described (2). This low seroprevalence is here observed in a population of treated patients with a favourable clinical evolution and outcome in most of these patients. In contrast, we identified that patients with severe disease developed a serological response in most cases (and all patients who died) that was characterised by high levels of IgG; in agreement with previous reports that antibody levels were higher after a severe and critical infection than after a mild infection (14-16). An immediate antibody response was observed in severe cases while it appeared later in mild cases (15, 16). On the other hand, an analysis of patients with mild symptoms of COVID-19 showed that SARS-CoV-2 can persist in patients who developed specific IgG antibodies for a very long period of time, up to 35 days, whereas a patient who did not develop an IgG response cleared the virus after 46 days (17).

Thus, high antibody titres were associated with severe disease regardless of age, gender and comorbidities, and there was no correlation between an early adaptive humoral response and improved clinical outcome (14). These results therefore call into question the much hoped-for role for serotherapy in SARS-CoV-2 infection. The use of convalescent plasma with high levels of neutralising antibodies planned at the onset of the pandemic for the treatment of severe COVID-19 infections may not be an effective treatment option (18-20).

Detecting anti-SARS CoV-2 antibodies is useful as a marker associated with COVID-19 severity. Serology also assesses exposure to the virus, at the individual level for middle-long term medical monitoring of the patients; and at the population level for monitoring the circulation of the virus, as it is one of the markers contributing to assessing the effectiveness of countermeasures.

## Data Availability

All data in the manuscript

## ACKNOWLEDGEMENTS

The authors acknowledge the contribution of the technical staff of the IHU Méditerranée Infection Laboratory. This work was supported by IHU Méditerranée Infection, Marseille, France.

